# ‘We need to feel that the Government value our lives’: Examining Irish women’s experience of cervical screening services

**DOI:** 10.1101/2024.08.23.24312417

**Authors:** Royanne McGregor, Sarah Foley

## Abstract

Cervical cancer is one of the top three cancers diagnosed in women globally. When women have access to a testing programme, abnormal cells can be detected to prevent the development of cancer. Research to date indicates that social and cultural barriers are the top two barriers in accessing cervical screening. In Ireland there may be a lack of trust in the National Cervical Screening programme due to previous mishandling of tests, and media coverage of the impacts of inaccurate screening results in Ireland. To understand impact of the health scandal on women’s trust in cervical screening in Ireland, nine Irish women ages between 25-65 were interviewed. Data was thematically analysed using a feminist perspective to centre the women’s voices in making sense of their appraisal and engagement with services. The data analysis resulted in four themes: Personal Reflections on Systemic Failure; Collective Concern and Blame; Decision Making Influencers and Rebuilding Knowledge and Trust. This analysis conveys distrust in the Irish Cervical Screening programme, feelings of anger, and a sense of neglect from the services. Irish women now place their trust in each other, the experience of their friends, families, their local GP. We discuss these finding to explore how Irish women have reinterpreted the narrative over the screening test in Ireland, and the potential to decrease concern around the topic by incorporating this experience into official narratives. This scandal heightened existing distrust and concern for the quality of women’s screening services, and therefore has global relevance which can be applied to screening services more generally.

## Introduction

Cervical cancer is ranked in the top three cancers present in women under the age of 45 (1,2) and accounted for 341,831 deaths across the globe in 2020 (3). Despite the high global mortality rates, cervical cancer is attributed to being one of the most preventable cancers and has seen a rapid decline in cervical cancer related deaths in Western countries (4). These reductions have been attributed to greater access to preventative vaccines, and higher levels of awareness of the importance of screening among women (5). In recent years, testing for Human Papillomavirus Virus (HPV) during screening has become common practice. HPV is a sexually transmitted disease (STD) and ranks as the most common STD worldwide (6). Persistent HPV can cause changes to the cells of the cervix, which may develop into cervical cancer (7). HPV cervical screening has shown higher accuracy levels and lower variability compared to cytology-based screening and is now the recommended screening method across Europe (8).

Despite the well-researched and referenced benefits of cervical screening, screening programmes and uptake varies considerably across the globe (1,2). Previous research has reported societal and psychological barriers to uptake. Some commonly reported barriers are fear, embarrassment, a belief of having a low risk of cancer (9), the test taking too long, or the test being painful (10). Inadequate knowledge and lack of awareness of cervical cancer is a major universal barrier for testing (11). A systematic review of experiences of the screening programmes internationally detailed the common reporting of the procedure as a physical and emotional ‘big deal’ (12). Furthermore, while the HPV screening method has increased accuracy and is highly valuable in detection, it has presented additional barriers. Literature has reported on stigma associated with both the HPV vaccine and screening across cultures, due to its association with sexual activity. Women have reported being worried about the perception they are promiscuous, or lacking in loyalty to their partner, resulting in a reluctance in screening (13,14). In contrast, women who are aware of symptoms, signs, causes, risk factors and prevention methods of cervical cancer are at an increased probability of attending screening (10). Additionally, age has been reported as a determining factor in test uptake, as women who had entered menopause, already had HPV, or were no longer of childbearing age were less likely feel the need for a test (15,16). The international literature demonstrates complex social, physical, and psychological barriers to engaging with screening services, despite the success of screening programmes in early detection of HPV. The Irish context presents further complexity in relation to engagement with the service, which highlights the impact of intense public scrutiny on trust in screening services. We introduce the Irish Health Services context below to situate our study.

### Cervical Screening in Ireland

In Ireland, the national cervical screening programme (CervicalCheck) offers free cervical smear testing to all Irish women aged between 25-65. Approximately 270,000 women are called per year for screening, with high uptake of 80% reported (7). Despite the efforts of national screening services, the benefits of screening have been overshadowed by public awareness of previous inaccurate testing. In 2018, Vicky Phelan, who had had received a terminal diagnosis was awarded a settlement of €2.5 million against a US laboratory that had inaccurately missed abnormalities in cervical screening she had undertaken. She was not told about the previous inaccurate testing at the time of her diagnosis. The case garnered national and international attention and, in a follow up investigation, it was reported that over 221 Irish women had received a false negative smear test result (17). The Scally Report, stated that apologies were only made to 43 women, meaning the majority were not told they received an incorrect cervical smear result (17).

While public figures vary in terms of the women impacted, it is estimated that up to 30 women have died to date as a result of misdiagnosis (18). These events are commonly referred to as the ‘CervicalCheck cancer scandal’ in the national and international press and saw high-profile coverage of women and their families who came forward to share their experience (18,19). High profile media coverage of the trials, advocacy and deaths of women including Vickey Phelan, Lynsey Bennett and Emma Mhic Mhathúna have highlighted the personal loss and suffering caused by the scandal (20). Further, advocacy groups for the women impacted have reported a sense of ‘deep mistrust’ in women’s health services in Ireland and calls for reform. An interview study with women who received inaccurate results examined the psychological impact of receiving this news, detailing a prevailing sense of loss and injustice in the aftermath (21). Despite reassurance in a follow up report by Dr. Gabrial Scally that women should be confident in the legitimacy of the service (17) the impact of this publicity on individual experiences of the screening programme remains under-examined.

Previous qualitative research into the impact of ‘health scandals’ for the provision of women’s health services, highlights the impact of media coverage of cases on medical injustice relating to the oral contraceptive pill (22,23). This literature demonstrates the intersection of politics, power and personal health, where women question the quality of healthcare provision provided to them, within a medical system that has traditional neglected the unique health needs of women Previous research on the uptake of the screening services in Ireland has examined the behavioural influences on cervical screening (24). Similar to international research, Irish women report a perception the test will be uncomfortable, with 67% indicated the test was “Unpleasant” (25). A study investigating behavioural influencers on cervical screening uptake among Irish women eligible for screening (26) highlighted the impact of social and environmental factors, age, knowledge, physical/psychological capacity, and perceptual and practical influencers (26). For example, older women often felt there was no need for a test if they were no longer sexually active. This study also noted the role of personal motivation and the impact of health professionals (26). A qualitative study conducted at the height of public attention of the scandal noted a ‘loss of trust, faith and confidence in the screening programme’ (27). Participants mentioned how the increased publicity could have positive effects such as improved services for the future and an increase in awareness of cervical screening (27). Taken together, these studies suggest considerable reform and discussion which centres the concerns of women is needed to rebuild trust in the screening services post- 2018. The Irish CervicalCheck, and the stakeholders involved in the service are faced with the complex challenge for restoring confidence in the screening programme, while also examining the lasting impact of the publicity surrounding the scandal on women’s decision-making to present for screening. Feminist Health Psychology literature conceptualises reproductive health as ‘entwined with social, cultural, and political practices’(28)p. 2) that reflect women’s place in society and their ability to exercise control over their reproductive health. The response to the Cervical Screening Scandal in Ireland is situated within a history of restricted access to contraception and abortion services, as well as historical mistreatment of women under the care of the State (29). Recent years have seen process through legislative changes, access to healthcare, and additional funding for a Women’s Health Action Plan (30). While this changing healthcare landscape is welcomed, it is important to examine how women engage with social and political narratives on women’s health and how it pertains their trust and engagement in cervical screening services. As noted above, there are many psycho-social barriers facing women when engaging with cervical screening. The Irish experience has additional socio-political factors that warrant further examination and response (31).

Given the widespread reporting of the fallout after the Cervical Screening Service scandal in Ireland, and the importance of screening services for early detection of abnormal cells, it is important to examine the prevailing perceptions of the Cervical Screening Services. As mentioned above, we employ a feminist health psychology (28,32) perspective to ensure women’s voices are centred in the response to this scandal. In the rest of this paper, we outline our study design, present findings from a feminist thematic analysis of nine semi-structured interviews with Irish women, and discuss these findings in light of international research on feminist-informed health service provision.

### Study Aims

This study aims to the examine the perspectives of a sample of Irish women who engage with Cervical Screening Services in the Republic of Ireland. Qualitative semi-structed interviews were conducted with nine women and analysed using feminist reflexive thematic analysis.

The study will seek to answer the following research questions:

- What are the current perspectives of women attending Cervical Cancer Screening in Ireland?
- How is the experience of engaging with the cervical screening programme constructed in relation to their perceptions of the CervicalCheck Scandal, and wider women’s health services in Ireland?

## Methods

### Participant Demographics

Data was obtained through interviews with nine women about their experience of obtaining a cervical smear test in Ireland in the period between 01/12/2020 and 30/04/2021. In Ireland, a woman is required to be aged between 25-65 in order to be eligible for the national cervical screening programme, therefore only females aged between 25-65 have been recruited for interviews. Gender and presence of a cervix have been used as exclusion criteria. In agreement with the ethical approval obtained, women who received results indicating they required further testing or follow up procedures after their latest smear test have been excluded due to differences in treatment. Convenience sampling strategies were used to recruit participants. Recruitment materials were shared across the social media and professional networks of the co-authors, and nine women came forward to participate. As noted in our findings below, the women who participated often referenced their concerns for others as a motivation to come forward for this study, and wanted to lend their voice to research on women’s health. Table 1 presents details of the participants pseudonyms and demographic details.

**Table 1:** Outline of participants.

| Pseudonym | Age Category | Number of smear tests |
| --- | --- | --- |
| Lola | 20-25 | 0 |
| Olive | 26-31 | 1 |
| Sally | 41-46 | +3 |
| Niamh | 36-40 | 2 |
| Emily | 47-52 | +3 |
| Eva | 47-52 | 3 |
| Aoife | 26-31 | 1 |
| Ray | 36-40 | 2 |
| Lilly | 26-31 | 2 |

### Design

Nine interviews were conducted by the first author, using a semi-structured interview style with questions focusing on reasoning behind obtaining a cervical smear test, personal attitudes to the test, motivators, demotivators, influence of the media. Interviews lasted on average 50 minutes and were conducted via video conference call due to local Covid-19 restrictions. The semi-structured format allows the participant to share new ideas and lead the conversation whilst the interviewer can ensure all the questions are answered. An interview schedule was drafted after a comprehensive literature review on the topic to explore women’s perspectives on their own experience of engaging with Cervical Screening. Ethical approval for this study was received from the University Ethics Committee. A pilot interview was conducted to ensure the questions were communicated well to participants. All participants provided written informed consent prior to taking part in the study. Participants were not known to either author prior to the interviews. As cervical smear testing can be a sensitive topic this format was chosen for its advantages in enabling participants to explore their thoughts, feelings, and beliefs, and delve deeper into personal and sensitive areas where they feel comfortable (33).

### Data Analysis

This study was informed by scholarship from feminist health psychology, which is concerned with the lives and experience of women and girls in society, whilst examining the impact gender has on power and influence in society (34). Feminist psychology roots itself in the understanding of women’s power and privilege in society, challenging the disadvantaged status of women and advocating on their behalf (35). The aim of our feminist reflexive thematic analysis is two-fold. We wanted to 1) explore the lived experience of women engaging with Irish cervical screening process, and 2) examine how socially available rhetoric’s based on the public reporting of the CervicalCheck scandal are used by Irish women to create individual positions. Based on these aims, we applied a reflexive thematic analysis (36), which takes a feminist perspective (37) Previous literature adopting a feminist thematic analysis emphasises its strategic use for centring the voices of women (38) while also informing change in services (39). Thus, in line with previous literature adopting a feminist approach to thematic analysis (40), we foregrounded the lived experience and idiographic details of the participants, to acknowledge the personhood of women engaging with larger health care services, and encourage critical response to the dominant power structures that have traditionally undervalued this perspective.

Once the interviews had been conducted, they were transcribed for data analysis.

Following the established steps of Reflexive Thematic Analysis, this process involved familiarisation with the data with repeated readings followed by line-by-line coding of each transcript (36). Feminist thematic analysis falls within the BigQ paradigm, which resists attempts to reinforce positivist norms within qualitative research (38). With this in mind, we coded each transcript individually, and then engaged in a process of close comparison of the analysed data to consider the depth of findings within the coded interviews, and final themes.

### Researcher positionality

The position of the researcher in relation to their qualitative research is a much-debated topic, with advantages and disadvantages for both insider and outsider research (41). As psychology researchers, we adopted a feminist poststructuralism (37) perspective to research design, which emphasises the importance of centring women’s voices, and examining language of resistance of dominant power structures. Feminist approaches to qualitative research also call for embodied reflexivity and aims to build a reciprocal relationship with the participants, sharing knowledge, respect, and empathy. The researcher takes the position that they are learning from the researched, valuing their input on the given topic rather than exploiting their data (42). This process involves the researcher embodying their self in the research (43) and learning from the participants. Feminist approaches also call for high levels of reflexivity in relation to researcher positionality. In conducting this research, both authors considered their own positionality regarding the engagement with women’s health screening services in Ireland and reflected on the shared experiences outlined within the data. They discussed how this influenced their data analysis and were careful to report on varied views presented in the data.

## Results

Our Analysis comprises of four themes: Personal Reflections on Systemic Failure Collective Concern and Blame; Decision Making Influencers and Rebuilding Knowledge and Trust. These themes detail the personal reflections on the cervical screening scandal and mechanisms employed to move past negative appraisals of the service. The findings outline subjective reflections on how the scandal has lasting impact on women, and their shared concern for how to move on post-scandal. They further offer insights into the potential ways forward for the screening service and emphasise the importance of relational trust in rebuilding confidence. We present the analysis below.

### Personal Reflections on Systemic Failure

The women we interviewed made sense of their experience within wider reporting of the 2018 CervicalCheck scandal. This was used to make sense of their own experience of the service. Participants conveyed little confidence in the smear testing procedure, portraying a sense of fear or hesitancy in taking up the screening and distrust in the result of the test.

Despite this mistrust, they continued to engage in the services. For example, a sense of distrust in the service in relation to prominent court cases resulting from inaccurate testing is conveyed:

> “All of them court cases…but it also makes me think, do I want to go for a private smear because the public system has had so many failures, like in a good way they are influencing you but in a bad way as well, where you are questioning everything that has come back.” Aoife

This example highlights the impact of hearing about multiple high profile court cases, in which women or their family members took legal action against the state and the laboratory that carried out the testing. The examples drawn on demonstrate conflicted feelings between taking up the test and acknowledging the failings of the service in the past, showing how these two concerns are at odds with each other. Elsewhere in the data, Eva justifies her mistrust in the service:

> “I think it’s disgraceful obviously because so many people have died from incorrect answers and incorrect results, and I think people are always nervous about that now.”

This example outlines a common declaration across the dataset in which the system is constructed as ‘disgraceful’ ‘poor’, or ‘a failure’. The participant conveys an idea that all women are similarly nervous, implying a belief that her worry is not unique, but a shared concern. Eva further evaluates the screening service based on reporting of the misdiagnosis of women rather than her own experience of the service.

> “Obviously poor, having seen the way women have been treated and having to drag their families through courts and everything and dying in the process, it is deplorable.”

This example draws on violent imagery of women being ‘dragged’ through the court system in a bid to seek justice, and further instances where women have died. A sense of luck was attributed to a positive screening experience:

> “There is that slight element of luck the results may not be accurate but it’s better than nothing.” Lilly

This belief that the tests are better than nothing, conveys a reluctant engagement with services. In contrast, another participant conveys an ease in engaging with the screening service:

"I can honestly say, I’m in and out and it’s not a bother, it’s not a bother, I don’t even give it a second thought". Emily

This quote is a contrasting but important insight into an alternative evaluation of the service. This theme highlights how women have interpreted the high-profile coverage of the misdiagnoses of other women in their overall appraisal of the service. This negative appraisal of the services as a systematic failure, often overrides their personal experience with the service.

### Collective concern and blame

The previous theme detailed how the women we interviewed constructed and justified their personal mistrust of the screening services. Throughout the data, wider concern for how other women were impacted by the cervical smear scandal was also prevalent. For example, concern regarding the consequential lower uptake in screening services is raised:

> “I am concerned that the scandals are putting people off getting them done rather than showing the problems, like what could happen if you don’t keep on top of it.” Ray

This quote shows concern regarding the consequences of the scandals on other women’s uptake of the screening service. While regularly screening is conveyed as important, there is a concern that the anger towards scandal has overshadowed the potential health consequences of not attending screening. Within the data, an alternative interpretation of how the scandal could be interpreted going forward:

> “God help the poor women who have lost their lives because of misread smears… like they are the key reasons why you need to go and get your smear.” Aoife

The above example offers a different response to the public awareness of the scandal. For Aoife, the women directly impacted are viewed as people to honour going forward and a source of motivation to continue returning for screening. In contrast, the government and associated health bodies are called out in their handling of the scandal. Elsewhere in the data, Eva constructs the Government as central actors in reshaping the existing narrative regarding trust in the service:

> “They made a mistake, and they should just fess up and we need guarantees that this will not happen again in the future, we need to feel that the Government value our lives.” Eva

This attributes the mistakes made in the scandal to the government, thereby placing a responsibility on them to reignite trust in the screening services, and further conveys the importance of mentioning lives are at stake.

The errors of the national screening programme have been broadcasted widely across Irish media. Many participants questioned the wider negative impact of the reporting of the fallout of the scandal. The impact of this on women who hear this coverage is detailed:

> “The only time you ever hear about it when it’s in the news is when the HSE have cocked up the results. It’s literally all doom and gloom surrounding it, it’s never positive.”

The above quote conveys the personal impact of hearing only negative coverage of the service by the media. Olive suggests the media presented a one-sided reporting on the screening programme, which contributes to continues negative appraisal of the service. This implies that the media have a role to play in the collective sense-making surrounding the cervical check services. In contrast, Sally detailed the benefits of the media coverage:

> “While you think it would never happen to you, it’s helpful having it in the media and having that in front of your mind reminding you why it’s important to get these tests.”

This sentiment echoes previous examples which seek out meaning from the wider reporting of the scandal. Participants draw on the worst-case scenarios and the impact they have when presented by the media to the public. This theme conveys the concern over how the scandal was reported on, and the impact this continued coverage has on women’s decisions to go for their smear test. Further, responsibility in moving forward is attributed to the government, the media as well as individuals themselves.

### Decision making influencers

While social and cultural narratives around are drawn on to justify women’s (lack of) trust and confidence in the screening service, decisions regarding uptake of the test were also attributed personal experiences with close family, friends and health care providers. This theme details the how women reflect on lived experience and seek out health information when deciding to take up the screening invitation.

The accessibility of the information available on the national cervical screening programme plays a vital role in helping women to make an informed choice suited to their needs. Participants outlined the importance of visibility of this information to counteract the wider reporting of the mistreatment of women, and the central role of the GP in providing information:

> "A lot of people will listen to their GP, you know". Emily

> “I think it should be more readily available. You know how they do campaigns like they put stuff on the back of toilet doors, like you know public toilets and things like that and sponsored ads on Facebook or something and I think when you go to the doctor it should be like lots of pamphlets there about it.” Eva

Despite high-profile campaigns across the media, there is a sense in the data that information needed to be more visible in social settings, including digital media. Emily suggests a sense of fear surrounding the test remains:

> “Well, a barrier really is fear, fear of the whole test, fear of what it involves, fear of… ‘I don’t know exactly what is about to happen.’”

As well as reliable information as a resource to base decisions on, the embodied experience of the test itself impacted women’s decisions to return for further screening. For example, Ray described her physical and psychological discomfort during the procedure, while Eva further described her experience of screening as ‘invasive’:

> “I was feeling a little bit vulnerable and a bit like yucky.” Ray

> “It is invasive, and I don’t like that.” Eva

The nature of the screening (obtaining swabs of the cervices) was uncomfortable for Ray and Eva. They both detailed the experience of attending for screening as evoking a sense of vulnerability. Perhaps in response to this sense of vulnerability, Ray conveys her preference for a more ‘friendly’ image of the service itself:

> “Obviously it is medical, but it is kind of not friendly in a way, probably could do with a bit of brand work.”

Participants expressed that a more ‘patient-friendly messaging’ would be welcomed, suggesting the service needs a re-brand:

> “A whole rebranding of it, definitely because it is so outdated, the letters are outdated, it is so outdated it so not the point, they sent hundreds of leaflets that aren’t even that relevant and they stress you out.*”* Aoife

This example acknowledges both the efforts of the national screening services to reach women but considers this process of sending out leaflets as outdated. This call for a rebrand implies a willingness to continue to engage with the service, despite the distrust outlined in previous themes. Overall, this theme conveys the potential benefits of rebranding the official communications of the national screening service as a means of rebuilding trust in the programme. The final theme further details the paths forward women have devised in order to continue to make use of the screening service.

### Rebuilding Knowledge and Trust

Due to the mistrust in the institutions responsible for the screening service, the women we interviewed took on the responsibility to empower themselves and ensure accurate information and encouragement is passed to others in their social networks. **Participants** also conveyed the key role of GPs and practice nurses in encouraging women to be tested, conveying the potential of relational trust and knowledge exchange. There was a sense that gaining more knowledge about the process allowed for more confidence in seeking out the screening.

For example, Eva outlines how she became more informed about the test through self-guided education:

> "I didn’t know they were now testing for the HPV virus instead of testing for abnormal cells, I only found that out by accident and that was through a webinar I did."

Similarly, another participant outlines how being aware of what is involved in the screening process helps her feel comfortable during the test:

> “I was nervous going in but the nurse I normally go to she would talk you through everything she goes through the whole process and that is kind of comforting.” Niamh

As well as educating themselves, the women we interviewed conveyed their sense of responsibility towards others. Lola described how she would encourage her family to get tested as she wants to make sure they are safe and looked after for peace of mind for herself:

> “I would want people in my family to get tested just because we hear about all the negative things that have happened its actually so scary to think like it could cause death.”

The willingness to be open and discuss the topic of cervical smear testing has been attributed by the participants to encourage others to get tested but also reduce the stigma and societal collective embarrassment. For example:

> “I kinda thought it was important to share, that but I did think a few people did book their appointments based on that conversation being so open.” Ray

While participants convey the opportunity to share encouraging accounts of the screening experience, our participants also presented a concern that the traditional lack of openness has affected women’s perceptions of the test and permanently challenge the narrative around smear testing in Ireland. For example:

> “I just feel it is a really taboo subject when it shouldn’t be, it should just be the same as any sort of check-up really, and the fact that it’s so taboo I think people don’t want to talk about it, I don’t know why but it’s like making it the secretive horrible thing.” Olive

Here Olive compares the test to a health check to highlight the difference in attitude between the smear test and other health checks.

Moving on from the Cervical Screening Scandal requires the appropriate honouring of the women who spoke out after receiving inaccurate results:

> “As terrible and as sad and as tragic it is for people to get the wrong results and it may be too late for these people, but I think them speaking out helps to save others, and it helps them take action, definitely it has helped me.” Sally

This theme presents the importance of trusted others in reestablishing trust in the services. However, a sense of ‘too little too late’ is evident in the final illustrative quote, conveying the complex emotions that remain regarding these services. In the final section, we discuss the implications of this study for healthcare provision going forward.

## Discussion

This Feminist Reflexive Thematic Analysis examined the experience of engaging with cervical smear screening in Ireland. Our analysis presents four themes: Personal Reflections on Systemic Failure; Collective Concern and Blame; Decision Making Influencers and Rebuilding Knowledge and Trust. The accounts of Irish women convey anger towards the failure of the services, personal fear for themselves and others engaging with services going forward, and important considerations for rebuilding trust in the screening programme.

Despite the continued engagement with the screening programme reported by our participants, the high-profile coverage relating to accounts of the 221 women who had received incorrect smear test results (17) is central to constructing an understanding of the overall service. This sense of fear and mistrust echoes previous literature in the Irish context, implying a lasting cultural and personal impact from the fallout for women’s health initiatives (21,27). In the face of this, there was also a sense of individual commitment to their health as well as a concern for other women and people requiring cervical screening that presents opportunity for rebuilding trust and continued engagement in the service. Furthermore, the breakdown in trust experienced as a result of the CervicalCheck scandal has implications for designing trusted services globally. In the remainder of this section, we discuss the implications of these qualitative findings for cervical screening services, informed by feminist health psychology literature (28).

Research to date on the fallout of the cervical screening scandal has noted considerable impact on the women directly involved in receiving inaccurate results and their families (44). Our analysis provides further insight into how these high-profile cases have been interpreted to become central to the personal understandings of the service. This builds on previous literature outlining growing mistrust in the services (27) and provides insight into the mean-making process women have gone through in recent years. Our findings highlight how the social narratives regarding the mistreatment of women in Ireland has evolved to incorporate the cervical screening scandal. This understanding has health implications, as women shared both concern for themselves and others who may be reluctant to engage with services. With the increases in distrust in official medical messaging, women in this study detailed a strong reliance on social relations as a site for rebuilding trust, where women were trusting each and their own GP rather than the national cervical screening programmes promotional advertisements and leaflets. Many felt the need to advocate to increase uptake of cervical screening within their social networks, including one woman from this who study posted her screening procedure on social media while. Despite personal positive experiences with the screening service, there was a sense that a failure to any women evoked a personal, emotional response. For many, the stories of the women who received inaccurate testing had taken on symbolic meaning, and represented wider evidence of the treatment of women in the Irish health care sector. This presents important insights into the connections made by our participants between personal experience and wider social knowledge on the value of women’s health in society (28).

The striking concern for others evident in our data set implies an opportunity to further centre women’s voices in the messaging and information on screening, including their fears. Recent research by Klonteig et al. (45) has adopted a co-design approach to cervical screening services, and further highlighted opportunities for digital tools to support the healthcare interaction. This expands calls in our data for ‘rebranding’ and points to the role of digital messaging in reaching women. Further it is an opportunity to ensure women are informed about the risks and benefits of screening through media campaigns, given the emphasis in our analysis on the role of public health campaigns and media reporting in shaping personal evaluation of the service (46).

The process of obtaining a cervical smear test was described as “Yucky” “Invasive” and “uncomfortable”. However, it is worth noting many participants were comfortable with the process, especially when they were supported by the Health Care Professional conducting the screening test. Constructions of the experience as invasive and uncomfortable draw further attention to the embodied experience, and the role of the practitioner in mitigating these experiences at a facility level. The construction of self-stigma and discomfort in our analysis demonstrates wider discomfort with embodied expressions of gender, and the ‘leaky’ bodies of women (47). At the time of data collection, self-screening is not available in the Irish services, but is becoming a more viable option for screening internationally (48,49). The opportunity to self-administer the test is an opportunity to acknowledge the importance of choice in women’s reproductive healthcare and acknowledges individual variation in women’s relationships with their bodies and ability to exercise choice. This aligns with feminist health literature that calls for acknowledgement of the treatment of women’s bodies in society, and the need for greater autonomy and choice in reproductive healthcare (50).

The Irish government were frequently called out as being responsible for the mistreatment of Irish women in the CervicalCheck Scandal. Further investment in women’s health initiatives at a government level is a welcomed step forward in increasing access to health care, but has an important role creating visible social understandings of the value of women’s health in society. For example, there was a strong emotional response in our data towards the women who have died as a result of inaccurate testing. Moving forward, it is important for the social and official narrative on cervical screening continues to acknowledge these women as central to the narrative surrounding cervical cancer in Ireland. Recent examples of welcomed initiatives include The Laura Brennan HPV Vaccine Catch Up Programme, which was launched in 2023 to honour an advocate who publicly campaigned for uptake in the HPV Vaccine before her untimely death aged 26 (51). While these examples are of national significance in Ireland, they convey a more universal implication as to how women interpreted and make sense of the treatment of other women accessing healthcare, and the personal process involved in making sense of political, social and embodied knowledge regarding reproductive health. The overwhelming focus on the CervicalCheck Scandal as a source of knowledge also suggests the need for enhanced investment in educational interventions. Previous research has reported educational interventions as highly effective in increasing cervical screening uptake, and may include media-based messaging, community outreach by healthcare providers and group discussions (52). Through their own advocacy work, the women in this study have already undergone this process privately where they have taken to their own social media and friend groups to support each other. Some participants further noted finding information ‘by accident’ through conversations or online resources.

Increased investment in educational interventions have important role to play in ensuring information that is shared is from valid, evidence-based sources. This is especially important when there has been a breakdown in trust in the official services.

## Strengths and Limitations

This study contributes an in-depth perspective on cervical screening experiences in the Irish context. While our sample size of nine interviews was limited to Irish women, striking similarities in the accounts offer important insight into how media-led national discussions on screening and co-opted into personal accounts and experience. All the participants were Irish Nationals living in Ireland, which brings up questions regarding the experience of non-Irish nationals in engaging with both the services, and wider experiences on the scandal. The women who came forward for interview were generally concerned for the future of services and how to keep other women committed to routine screening. In future research, it would be important to recruit participants who have disengaged from attending services, as well as women who required follow up procedures after the initial screening.

Further, given the participants who came forward had engaged in self-directed research on the process, future research should engage with women who may have less access to digital health information.

## Conclusion

The experience of obtaining a cervical smear test in Ireland is at the intersection of embodied knowledge, cultural understandings of women and healthcare provision. Despite the benefits of regular cervical screening, the women in our study constructed accounts of fear and a lack of confidence in the cervical screening services. This distrust has resulted in a sense of reluctant compliance with screening services. The Reflexive Thematic Analysis conveys the importance of repositioning women’s experience as central to the shared information on screening and calls for a commitment to strong messaging and investment in the service from official stakeholders.

## Data Availability

This qualitative data contains potentially identifying or sensitive information. Parts of the data sets can be made available upon request.

